# Uncertainty in serious illness: A national interdisciplinary consensus exercise to identify clinical research priorities

**DOI:** 10.1101/2023.07.21.23293007

**Authors:** Simon N Etkind, Stephen Barclay, Anna Spathis, Sarah A Hopkins, Ben Bowers, Jonathan Koffman

## Abstract

**Background:** Serious illness is characterised by uncertainty, particularly in older age groups. Uncertainty may be experienced by patients, family carers, and health professionals about a broad variety of issues. There are many evidence gaps regarding the experience and management of uncertainty.

**Aim:** We aimed to identify priority research areas concerning uncertainty in serious illness, to ensure that future research better meets the needs of those affected by uncertainty and reduce research inefficiencies.

**Methods:** Rapid prioritisation workshop comprising five focus groups to identify research areas, followed by a ranking exercise to prioritise them. Participants were healthcare professionals caring for those with serious illnesses including geriatrics, palliative care, intensive care; researchers; patient/carer representatives, and policymakers. Descriptive analysis of ranking data and qualitative framework analysis of focus group transcripts was undertaken.

**Results:** Thirty-four participants took part; 67% female, mean age 47 (range 33 – 67). The highest priority was communication of uncertainty, ranked first by 15 participants (overall ranking score 1.59/3). Subsequent priorities were: 2) How to cope with uncertainty; 3) healthcare professional education/training; 4) Optimising clinical approaches to uncertainty; and 5) exploring in-depth experiences of uncertainty. Research related to optimally managing uncertainty was given higher priority than research focusing on experiences of uncertainty and its impact.

**Conclusions:** These co-produced, clinically-focused research priorities map out key evidence gaps concerning uncertainty in serious illness. Managing uncertainty is the most pressing issue, and researchers should prioritise how to optimally manage uncertainty in order to reduce distress, unlock decision paralysis and improve illness and care experience.

**Key points:**

- Uncertainty is ubiquitous and distressing in serious illness, and can paralyse decision making
- In this consensus exercise, stakeholders identified research priorities for uncertainty in serious illness
- Communication of uncertainty was the highest priority
- Participants prioritised research concerning managing uncertainty above research to understand experiences of uncertainty

## Background

Uncertainty is ubiquitous in serious illnesses across all health settings, especially when living with long- term conditions and frailty.[1–5] Encompassing “known unknowns”, uncertainty is characterised as an inadequate understanding, a sense of incomplete, ambiguous or unreliable information, and conflicting alternatives.[6, 7] It is inherently a complex concept and situations of uncertainty often result from several inter-related factors.[8]

Irrespective of its origin, uncertainty matters because when suppressed and ignored, it can profoundly negatively impact patients and their family.[9, 10] Older adults may be particularly affected as they commonly experience complex and unpredictable illness, associated with irreducible uncertainties. Uncertainty may precipitate extensive psychological and existential distress, potentially culminating in an experience of ‘Total Uncertainty’ which may threaten an individual’s sense-of-self.[11]

If uncertainty is not addressed it may impact patient safety, adverse events, healthcare interactions and relationships.[12, 13] One metric where this is recorded are complaints levelled at healthcare, particularly in hospital settings.[14] Uncertainty can also limit patient participation in decision-making, leading to ‘decision paralysis’,[15, 16] which may contribute to sub-optimal care and has repercussions for the allocation of scarce health resources, including hospital admissions and longer inpatient stays.[17, 18]

It is not just patients who are affected by uncertainty. Despite uncertainty in medicine dating back to Hippocrates, there exists a deeply rooted aversion to it in empirical medicine, where acknowledging uncertainty can have connotations of failure.[18, 19] If poorly tolerated by health professionals, uncertainty can adversely impact their confidence and competence, increasing the risk of moral injury, burnout and depression.[20–23] This was particularly evident during the Covid-19 pandemic.[24, 25]

Uncertainty in illness is not necessarily distressing; it may be appraised negatively, neutrally, or in some individuals, illnesses, and care circumstances, positively; for example holding onto prognostic uncertainty can in some circumstances enable people to retain hope.[26–28] The distress caused by uncertainty, and hence the block to decision making is therefore not inevitable; the negative impacts of uncertainty can be at least partially ameliorated if it is addressed and communicated sensitively.[29–31]

There are innumerable possible situations of uncertainty, each of which may have its own optimal approach. We still do not know how best to approach and address it in older patients living with long- term or life-threatening illnesses in a way that best supports the individual and those involved in their care;[32, 33] although previous work has explored how to support communication of uncertainty, [34] uncertainty management,[12, 32] and shared decision making.[35] We do know that there can be no one-size-fits-all approach to uncertainty, as it is experienced differently in different clinical contexts, by different individuals.[6, 36] Uncertainties in some contexts may be more distressing than others and may require different approaches.[37] Despite the importance of uncertainty for patients and clinicians alike there has, to date, been no attempt to prioritise the most pressing areas to focus applied research on to improve care.

We aimed to identify stakeholder priority research areas concerning uncertainty in serious illness, [38] to enable future research to more effectively meet the needs of those affected by uncertainty and to reduce research inefficiencies.[39, 40]

## Methods

### Design

National interdisciplinary one-day prioritisation workshop with a range of stakeholders: patient and carer representatives, clinicians, researchers, and policymakers. Reporting according to REPRISE guidance (see appendix).[41]

### Patient and public involvement

A Patient and public involvement group supported development of the study aims and methods; 3 public contributors participated in the workshop, contributing to focus groups and the ranking exercise, and commented on the findings.

### Participants and participant identification

Participants were clinicians, researchers, policymakers, people with lived experience of serious illness and their informal carers. Researchers and policymakers were eligible if they were interested in the area. Clinicians were from any profession or specialty with experience in providing care to people with serious illnesses. The workshop invitation was disseminated widely through clinical and research networks and social media, including: Applied Research Collaborative (ARC) East of England, UK uncertainty in serious illness specialist interest group, the UK-wide Community Nursing Research Forum, regional and national palliative care and gerontology contacts. Invitations were sent from December 2022, and registration was open until 27^th^ February 2023. The workshop was held on 28^th^ February 2023. Workshop attendees were informed of the research component in advance.

### Workshop process

The workshop drew on existing approaches to prioritisation, incorporating idea generation, consolidation, and ranking stages.[42] Written informed consent and self-reported demographic data were collected from workshop participants at the start of the day, including age, gender, ethnicity, participants’ status as a researcher, clinician, policymaker, or patient representative, and details of their field where relevant. To encourage debate, the workshop began with presentations on the “state of the science” concerning uncertainty in serious illness. Models of uncertainty and evidence gaps from relevant literature reviews were outlined.[11, 32, 34, 43, 44]

We then held focus groups with participants to explore their views on uncertainty and identify key areas for future research. This approach mirrored the idea generation stage of the nominal group technique and the first round of a Delphi process.[42, 45] The topic guide was developed by the research team and was informed by literature review: it focused on experiences of uncertainty, views on desired outcomes when addressing uncertainty, and ideas for research questions concerning uncertainty in serious illness. Focus groups were led by clinicians and researchers with expertise in facilitation. Conversations were audio recorded and a scribe within each group recorded the research questions identified by participants.

Research areas from the focus groups were collated into a summary list during the day. In the final session, this summary list was presented to participants who were invited to anonymously rank the top three areas in order of priority using the online ranking tool “Slido” (© 2023 Cisco Systems, Inc.). Items were presented in random order. See appendix for full workshop programme and topic guide.

### Data analysis

*Analysis of focus group lists:* The lists generated by focus groups were reviewed by two researchers (SE & JK) during the day, taking into account both researchers’ existing knowledge of evidence gaps and previously expressed areas for future research from literature reviews.(9, 28) The researchers combined the lists produced by each focus group by removing duplicates and arranging similar questions under “umbrella terms” to produce a single summary list of priority research areas for use in the subsequent ranking exercise.

*Analysis of ranking exercise data:* Individual-level ranking scores were exported from Slido. We analysed the scores descriptively and reported the number of participants ranking each item within their top three. We calculated an average ranking score by allocating points to each item as follows: the item ranked first received three points; the item ranked second received two points; the item ranked third received one point; all other items received zero points. The average ranking score was calculated by adding the points for each item and dividing by the number of participants. The maximum any item could score was three if every participant ranked it as their top priority and the minimum was zero if no participant ranked it in their top three.

*Analysis of focus group transcripts:* Following the workshop the focus group recordings were transcribed verbatim, anonymised and analysed using a framework approach.[46] This stage aimed to identify detailed research questions within the priority areas, as well as any additional areas discussed that were relevant for future research. We used the research priority areas identified during the workshop as a coding framework and one researcher (SE) coded text in the transcripts that described research questions or participants’ views about these areas. A second researcher (JK) independently reviewed one focus group transcript. Coding was reviewed, and where there were differences, these issues were reconsidered and debated by both researchers until consensus was achieved.[46] To avoid making unwarranted claims about patterns and regularities in the data, we examined and coded unusual or non- confirmatory views that did not fit easily into the original framework.[46] The framework was condensed, summarised and discussed with the wider research team to refine it. Anonymised excerpts from the transcripts are presented to illustrate themes and represent a range of views.

### Ethical approval

This study received approval from the University of Cambridge Psychology Research Ethics Committee [Reference:PRE.2022.125].

## Results

### Participant details

Thirty three participants took part in the focus groups, and 34 in the ranking survey, of whom 30 provided demographic information. The average age was 47 years (range 33 – 67), and 67% were female. 80% were of white ethnicity, 10% Asian, and 10% from mixed or multiple ethnic groups. 70% were clinicians, 43% researchers, 10% patient or carer representatives, and 7% policymakers (participants could state multiple roles). Of the clinician participants, twelve had a background in palliative care, three geriatrics, two nursing and one each of intensive care, general practice, psychology, and physiotherapy.

### Item generation and ranking

Five focus groups with six to seven participants in each were of 53 to 64 minutes’ duration. The groups generated 61 research questions, which we condensed to produce the 10 priority areas that were then ranked by participants (Table 1). Communication of uncertainty was the highest-ranked item, scored first by 15 participants. Participants ranked the next four priorities almost equally: coping with uncertainty; training health professionals; optimising clinical approaches to uncertainty; understanding in-depth experiences of uncertainty. The other areas received lower priority scores.

**Table 1.** 10 priority areas for future uncertainty research in serious illness (n = 34):

| Overall Ranking | Priority | Number of participants ranking item in each top 3 positions (%) |  |  | Percentage of participants ranking items in top three | Ranking score* |
| --- | --- | --- | --- | --- | --- | --- |
|  |  | #1 | #2 | #3 |  |  |
| 1 | Communication of uncertainty | 15 (44) | 3 (9) | 3 (9) | 62 | 1.59 |
| 2 | How to cope with uncertainty | 6 (18) | 5 (15) | 3 (9) | 41 | 0.91 |
| 3 | Education/training of healthcare professionals | 2 (6) | 7 (21) | 9 (26) | 53 | 0.85 |
| 4 | Optimising clinical approaches to uncertainty | 6 (18) | 2 (6) | 4 (12) | 35 | 0.76 |
| 5 | Understanding patient/carer experiences of uncertainty in depth | 2 (6) | 7 (26) | 5 (15) | 41 | 0.74 |
| 6 | Variation in experience/response to uncertainty between different individuals, groups, professions | 1 (3) | 4 (12) | 1 (3) | 18 | 0.35 |
| 7 | Explore positive aspects of uncertainty | 1 (3) | 1 (3) | 4 (12) | 18 | 0.26 |
| 8 | Impact of uncertainty on bereavement | 1 (3) | 2 (6) | 1 (3) | 12 | 0.24 |
| 9 | Uncertainty in specific conditions/clinical situations | 0 (0) | 3 (9) | 1 (3) | 12 | 0.21 |
| 10 | Factors associated with different uncertainty experiences | 0 (0) | 0 (0) | 1 (3) | 9 | 0.09 |

\*Calculated as follows: item ranked first receives 3 points; item ranked 2<sup>nd</sup> receives 2 points; item ranked 3<sup>rd</sup> receives 1 point; all other items receive 0 points. The total points for each item are added and divided by the number of participants.

### Detailed research priorities

We explored the 10 priority areas raised by workshop participants during qualitative analysis, and identified detailed sub-questions within each area (Table 2).

**Table 2.** Detailed research priorities from framework analysis*.

| Theme | Priority area | Detail of priority area |
| --- | --- | --- |
| Uncertainty management | Communication of uncertainty<br>(research on how best to communicate about uncertainty) | • Understand more about the detailed processes for uncertainty communication: who should be involved, optimum setting & timing, what phrases to use; how much uncertainty to share; how are such conversations received by patients<br>• How to communicate uncertainty openly whilst maintaining a trusting relationship?<br>• Interprofessional communication<br>• How to individualise communication of uncertainty to different settings, situations, individual characteristics?<br>• How does the communication of uncertainty affect decision-making and other outcomes? |
|  | How to cope with uncertainty<br>(how can individuals best be supported to cope with their uncertainties) | • How do health professionals cope when they feel uncertain and how do they achieve resilience/tolerance to it?<br>• How can patients and carers be supported to cope with their uncertainty and develop resilience?<br>• Who copes well and why?<br>• How to cope with the long-term impact of decisions made under conditions of uncertainty?<br>• What is the role of hope in coping with uncertainty? |
|  | Education/training of healthcare professionals (how can we train health professionals to approach uncertainty) | • How to teach uncertainty management to medical students, what would be the goals of such training?<br>• How to train HCPs at different levels to recognise/tolerate/hold their own uncertainty?<br>• How to transfer expertise from areas where uncertainty is well managed?<br>• Development of training interventions including psychological training |
|  | Optimising clinical approaches to uncertainty<br>(how can the uncertainty of others best be managed and addressed, what are optimal approaches) | • How to individualise the management of uncertainty depending on patient experiences, response to uncertainty, and how to identify how much uncertainty is tolerable to an individual?<br>• Who should 'hold' uncertainty and how to find a balance in terms of information sharing and decision-making<br>• How to maintain trust and manage expectations?<br>• How to gauge the readiness of people to discuss uncertainty and time conversations?<br>• How can technology be used to distil information and reduce/address uncertainty relating to complexity/information overload?<br>• What is the role of uncertainty management interventions and models of care to address uncertainty, and how effective are these?<br>• What are the key outcomes we should be aiming for when seeking to approach and manage uncertainty?<br>• What are the barriers to addressing uncertainty? |
|  |  | How to manage uncertainty as an MDT and maintain continuity of approach |
| Uncertainty experiences | Understanding patient/carers experiences of uncertainty in depth<br>(what are patient and carer experiences of uncertainty) | How does the experience of uncertainty affect decision-making?<br>What is the lived experience of uncertainty amongst patients and families (what is helpful when faced with uncertainty and what is harmful? How do the multiple layers of uncertainty interact within an individual's experience (mapping uncertainty)? Are the experiences/impacts of different types of uncertainty different? How do past life or healthcare experiences affect current uncertainty experiences.)?<br>How is uncertainty transmissible i.e. how can HCP uncertainty be picked up by patients and vice versa and what is the effect of this? |
|  | Variation in experience/ response to uncertainty<br>(how does the experience of uncertainty vary between different groups) | How does experience and response to uncertainty vary between different medical specialties and health professional groups?<br>How does experience and response to uncertainty change over time as the illness progresses.<br>How do individual characteristics e.g. culture, LGBTQ status, neurodiversity, age, affect experience and response to uncertainty?<br>How does career stage and knowledge/experience in a clinical role affect health professionals experience?<br>What are the long-term effects of uncertainty experiences in ITU on ITU survivors? |
|  | Explore positive aspects of uncertainty<br>(what aspects of uncertainty are positive and how can these be promoted) | What is the relationship between sharing uncertainty and maintaining hope?<br>Explore positive utility of uncertainty e.g. as a way to promote a quest for knowledge or changing ways of thinking<br>Explore why some people thrive with uncertainty |
|  | Impact of uncertainty on bereavement<br>(how does uncertainty in serious illness impact on bereavement experiences) | How does uncertainty in serious illness and at the end of life affect experiences and outcomes of bereavement? |
|  | Uncertainty in specific conditions/clinical situations<br>(the uncertainties that are experienced in a particular clinical situation such as in ITU) | How do different clinical situations change how uncertainty is experienced and responded to?<br>Are there situations where the expression of uncertainty is not appropriate?<br>How to make good decisions in the context of uncertainty in the intensive care unit?<br>How is uncertainty experienced in children with complex neuro-disability?<br>How to approach uncertainty in adults with neurodegenerative disease and uncertain illness trajectory?<br>How to approach uncertainty in frailty with uncertain prognosis? |
|  |  | • How to communicate uncertainty at transitions of care |
|  | Factors associated with different uncertainty experiences<br>(what factors are associated with different experiences of or responses to uncertainty.) | • What system factors underlie the challenges of uncertainty?<br>• What factors are associated with uncertainty experiences and tolerance e.g. age, life experience, culture?<br>• How did the COVID-19 pandemic affect experiences and response to uncertainty?<br>• How does trust in clinicians affect uncertainty experiences? |

The highest priority research areas were related to managing uncertainty, by communication, supporting individual coping mechanisms, or providing training. Participants particularly focused on how and when to have conversations about uncertainty, and how to individualise communication to promote psychological wellbeing:

> “Then there’s a really important research need around understanding more about how we communicate uncertainty and the consequences of how we communicate uncertainty and how we can talk about uncertainty in a way which is more patient-centred and supportive and considers sort of psychological wellbeing.” Social scientist, focus group 3.

Training of health professionals at varying stages was a recognised priority. This included training professionals on toleration of their uncertainty as well as how to address the uncertainty experienced by others. One participant highlighted that inadequate training of doctors to manage uncertainty has been a longstanding issue and queried the best timing of such training:

> “We’ve been inadequate at teaching them [medical students] how to actually manage this complexity for a long time, although you could argue that it’s quite difficult to teach until they really get into the nitty-gritty of practising.” Geriatrician, focus group 1

Participants recognised it is unlikely a single intervention can address all the nuanced multilevel aspects of uncertainty, but nevertheless, some felt interventions could play a role in addressing uncertainty, as long as they were situated in a broader societal context:

> “There are interventions that are being developed out there but there are no interventions that deal with all the different types and layers of uncertainty and they can’t by nature. And I think there is that kind of relational uncertainty, the kind of organisational uncertainties, there are uncertainties on macro, meso, micro levels and you can deal with one component, but you can’t deal with all the different components, and it’s how those interventions work within the broader societal context of uncertainty.” Occupational therapist, focus group 4

Older frail patients were seen as a priority for future research into how uncertainty can be managed. Participants noted a lack of knowledge about approaching uncertainty in the context of frailty:

> “For old and frail elderly there’s no support you know, for that anxiety, fear, so it’s how do we look at that, you know, and how do we maybe look at how we can support, supporting that frailty and older.” Professor of end-of-life care, focus group 5

Participants identified research questions concerning *experiences* of uncertainty, though these were generally given lower priority than questions relating to *managing* uncertainty. Participants identified illness contexts where uncertainty experiences could be explored further, including critical illness and frailty. They also acknowledged it is important to explore how the experience of uncertainty varies depending on perspective (patient, carer or health professional), or individual characteristics:

> “We’d want to look at different groups such as sort of learning disability, neurodiversity, sort of hard-to-reach areas whether deprivation, LGBTQ+, sort of that kind of differences you might get.” Palliative care consultant, focus group 1.

Participants identified potential positive impacts of uncertainty and its utility in certain situations. They suggested it was important to understand why some health professionals thrive when required to manage uncertainty, whereas others exhibit a lower tolerance. Participants noted the relationship between uncertainty and hope and reasoned that positive aspects of uncertainty should be explored further:

> “I wonder….whether there’s something about learning to cope with uncertainty or to tolerate uncertainty and whether people, patients and families can see that as a positive as well.” Bereavement practitioner, focus group 4

### Additional areas for future research

Some questions identified from focus group transcripts were not identified as research priority areas by participants. Additional areas included consideration of the legal and regulatory implications of uncertainty and its management, the link between uncertainty and patient safety, how to prepare the public for serious illness uncertainty, and the resource impacts of different levels of tolerance to uncertainty:

> “Have we studied the use of resources around uncertainty? Because I’m sure loads of tests are done completely unnecessarily because people just want to be sure.” Palliative care consultant, group 1

## Discussion

### Summary of findings

This study co-produced clinically-focused research priorities to address known evidence gaps concerning uncertainty in serious illness. Optimising communication of uncertainty was the top research priority. Research into managing uncertainty was considered higher priority than research investigating experiences of uncertainty.

### Discussion of main findings

Communicating uncertainty was the top priority for participants, reflecting key evidence gaps and recommendations in this field.[32, 47] In their narrative review of uncertainty communication, Simpkin et al identified a number of evidence gaps, including identifying individuals’ communication preferences and tailoring communication to those preferences.[34] The question of how to maintain hope whilst communicating uncertainty was noted as a priority; this has been explored in cancer care,[48] but remains a key question in other serious illnesses. Additionally, participants raised several sub-questions in terms of *how* to discuss uncertainty, reflecting the need for implementation-focused communication research.

After communication, participants prioritised other aspects of managing uncertainty: identifying how individuals can be supported to cope with their own uncertainty; investigating how we can optimise clinical approaches to uncertainty; understanding more about how to equip health professionals to deal with uncertainty through training. Though management of uncertainty has been recognised as a core component of medical training for decades, curricula still make limited reference to uncertainty, and filling this gap should be a priority.(2) Whilst we have an improving understanding of how physicians manage uncertainty,[31] the evidence base for other professional groups is very limited, yet nurses and allied healthcare professionals often lead the clinical care and support of older people and their families. To date, uncertainty management and communication interventions have had variable impact in serious illness,[32] and often prove challenging to evaluate.[33, 49] There is scope for further intervention development,[50] especially work that focuses on evaluation of uncertainty-focused interventions.

Whilst many of the questions identified by participants related to investigating experiences of uncertainty, these were usually considered lower priority, perhaps because much is already known about uncertainty experiences.[7, 51] For example, conceptual taxonomies of uncertainty are well developed,(6) and there have been evidence syntheses of experience in some specific areas e.g. multimorbidity.[11] However, there are still evidence gaps concerning how uncertainty affects individuals in other clinical contexts, in particular older patients living with frailty. Given the rapidly increasing complexity of the healthcare system and unpredictability of the frailty trajectory, this is an area that warrants urgent exploration.[34] Understanding more about the impacts of uncertainty on experiences of illness, care, and bereavement would enable us to develop interventions focused on the real-world problems uncertainty can cause.

### Strengths and limitations

The rapid prioritisation approach we used enabled a diverse group of interested individuals to generate and rank research priorities in a single day. Those ranking the priorities had spent the entire day considering uncertainty in serious illness and were well placed to express considered views when ranking the list presented to them. By identifying evidence gaps before the workshop and communicating these to participants during initial presentations, we were able to focus on areas where more research is needed and incorporate the key stages of a traditional prioritisation exercise. By additionally incorporating formal qualitative analysis we increased rigour and developed a robust priority list. This approach was a more feasible and pragmatic alternative to lengthier methodologies such as the Delphi process[52, 53]. However, the rapid nature of the prioritisation process meant there was limited time to condense the findings of the focus group discussions, which risked a loss of accuracy and ranking was limited to broad research areas. We ameliorated this by subsequent analysis of focus group transcripts, which enabled us to identify detailed research questions identified by participants, including areas that participants mentioned even when they weren’t identified as priorities at the time.

We incorporated patient and carer experiences, but most participants were healthcare professionals or researchers, thus these groups were not equally represented in the ranking process. Whilst not specifically excluded, social care professionals did not attend this workshop, which limits the findings to healthcare. We recruited a UK-wide sample, but this was not an international study, and future work should explore if these findings hold internationally, though literature from 17 countries reported consistent findings on a similar topic.[11] The anonymous nature of the ranking means we could not adjust for the background of participants when analysing ranking data. The largest group of clinical participants were from palliative care backgrounds which may have shaped their views; however, a broad range of health professionals and researchers were represented, and the degree of agreement, particularly regarding the top priority of communication suggests the findings represent true consensus.

## Conclusion

Through a rapid prioritisation workshop, we have identified 10 ranked priority areas for clinically focused research on uncertainty in serious illness. There was consensus that further research into managing uncertainty, particularly communication, was of higher priority than research to investigate experiences of uncertainty. Future targeted research could result in interventions to reduce the distress associated with uncertainty, unlock decision paralysis and improve illness and care experience.

## Data Availability

All relevant data from the ranking exercise is included within the manuscript and as a supplementary file. Due to ethical restrictions, the focus group transcripts cannot be made publicly available, however we have permission from the University of Cambridge Psychology Research Ethics Committee [Reference:PRE.2022.125] to make anonymised transcripts available to researchers who wish to undertake future related work. We will deposit the transcripts in the University of Cambridge Apollo data repository from the time of publication, with access controlled by the research team.

## Acknowledgements

Thank you to those who facilitated focus groups: Stephan Barclay, Jonathan Koffman, Anna Spathis, Ben Bowers, Sarah Hopkins, Debbie Critoph, Markus Schichtel, Ikumi Okamoto

We would also like to thank our patient and public involvement representatives: Roberta Lovick, Sarah Dixon, Rashmi Kumar

Many thanks to the workshop organiser Angela Harper & the professional services team at the Primary Care Unit, University of Cambridge for ensuring the smooth running of this study

Thank you to Zoe Fritz for your advice during drafting of the manuscript

## Contributions

Study design: SE, SB, AS, JK Securing funding: SE, SB

Data collection: SE, SB, AS, SAH, BB, JK Analysis: SE JK

Paper drafting: SE, SB, AS, SAH, BB, JK

Approval of final version: SE, SB, AS, SAH, BB, JK

